# Effect of Heat inactivation and bulk lysis on Real-Time Reverse Transcription PCR Detection of the SARS-COV-2: An Experimental Study

**DOI:** 10.1101/2022.04.04.22273334

**Authors:** Dereje Leta, Gadissa Gutema, Gebremedhin Gebremichael Hagos, Regasa Diriba, Gutema Bulti, Tolawak Sura, Desta Ayana, Dawit Chala, Boki Lenjiso, Jaleta Bulti, Saro Abdella, Habteyes Hailu Tola

## Abstract

The outbreak of the severe acute respiratory syndrome coronavirus - 2 has quickly turned into a global pandemic. Real-time reverse transcription Polymerase chain reaction is commonly used to diagnose as “gold standard”. Many coronaviruses are sensitive to heat and chemicals. Heat and chemical inactivation of samples is considered a possible method to reduce the risk of transmission, but the effect of heating and chemical treatment on the measurement of the virus is still unclear. Thus, this study aimed to investigate the effect of heat inactivation and chemical bulklysis on virus detection. The laboratory-based experimental study design was conducted in Ethiopian Public Health Institute from August to November 2020 on the samples referred to the laboratory for Coronavirus disease-19 testing. Tests were performed on eighty Nasopharyngeal/Oropharyngeal swab samples using the Abbott Real-time severe acute respiratory syndrome coronavirus - 2 assays, a test for the qualitative detection of virus in the sample. Data were analyzed and described by mean and standard deviation. Repeated measurement analysis of variance was used to assess the mean difference between the three temperatures and bulk lysis on viral detection. Post-hock analysis was employed to locate the place of significant differences. P-values less than 0.05 was used to declare statistical significance. About 6.2% (5/80) of samples were changed to negative results in heat inactivation at 60°C and 8.7% (7/80) of samples were changed to negative in heat inactivation at 100°C. The Cyclic threshold values of heat-inactivated samples (at 60°C, at 100°C, and bulk lysis) were significantly different from the temperature at 56°C. The efficacy of heat-inactivation varies greatly depending on temperature and duration. Therefore, local validation and verification of heat-inactivation are essential.

## Introduction

Coronavirus disease 2019 (COVID-19) is a newly emerged human infectious disease caused by severe acute respiratory syndrome coronavirus 2 (SARS-CoV-2) (1). SARS-COV-2 is a group IV-positive-sense, single-stranded Ribonucleic acid (ssRNA). Based on the rapid rate of increase in humans, the World Health Organization (WHO) classified the COVID-19 outbreak as a pandemic by the end of 2019 [1, 2]. SARS-CoV-2 infection is associated with COVID-19, which is characterized by severe respiratory distress, fever, and cough and high rates of mortality, especially in the elderly and those with underlying medical conditions [3]. Ethiopia is one of the 220 COVID-19 affected countries and territories, the first confirmed COVID-19 was reported on March 13, 2020, and the first wave peak of kurtosis of the epidemic was from August to September 2020. Since it has reached a critical point in March 2020, WHO declared that the world needs speedy and quick solutions to diagnose and tackle the further spread of COVID-19 [4, 5]. Amplification of the viral RNA by qualitative and quantitative real-time polymerase chain reaction (RT-PCR) is currently the gold standard procedure for diagnosis [1]. Molecular modeling studies demonstrate that, like SARS-COV, SARS-COV-2 is a membrane-bounded lipid bilayer containing a structural (M) and an envelope (E) membrane. This layer contains spike (S) glycoprotein that gives this virus family the characteristic “crown” shape [6, 7]. The SARS-CoV-2 RNA polymerase, E gene that codes for an envelope, nucleocapsid region, and the open reading frame (ORF) 1 are the most ideal amplification target. The N gene-based rRT-PCR assay was more sensitive than the ORF 1 assay for the detection of SARS-CoV-2 in clinical specimens [6]. The CDC rRT-PCR panel for detection of SARS-CoV-2 demonstrated high sensitivity and specificity for detecting RNA copies/reaction with no observed false-positive reaction [8], and it facilitates rapid detection of SARS-CoV-2 infections in humans.

These assays have proven to be valuable for rapid laboratory diagnosis and support, clinical management, and infection prevention and control of COVID-19 [9, 10].

Laboratory viral nucleic acid (NA) testing using RT-PCR assays is currently the “gold standard” for the diagnosis of COVID-19. However, the requirements of complex instruments and laboratory conditions, cumbersome experimental procedures, and longer detection times greatly hindered its large-scale applicability [1, 11].

Direct SARS-CoV-2 diagnosis is based on RNA detection by RT-PCR from nasopharyngeal- or throat-swab samples commonly containing high viral loads [4, 12]). Buffer-based NA extraction methods to obtain high-quality NA has not been developed primarily for the inactivation of infectious samples [13, 14]). Since extraction processes can produce aerosols, they must be performed in a Class 2 biosafety cabinet. However, automated NA extraction is often performed outside of the biosafety cabinet, which can lead to aerosol formation. To avoid this, a pre-inactivation step under appropriate biosafety conditions is an absolute requirement [13, 15, 16]. Accordingly, the extraction of viral RNA requires the first step of lysis or heat inactivation of the virus at different temperatures for different minutes [7, 17]. In our laboratory at the Ethiopian Public Health Institute (EPHI), we usually do heat inactivation for 30 minutes at 56°C. Despite the fact that a higher temperature takes a shorter period to lower infectivity [13], its impact on SARS-COV-2 detection has not been thoroughly examined. Furthermore, evidence indicated that infectivity of SARS-COV-2 is prevented after heating at various temperatures at different times. This is because the heat has an effect on SARS CoV-2 membrane protein [17, 18]. Even though it is known temperature influences the infectivity and virulence of the virus, still there is a lack of understanding of the molecular-level changes that are taking place in the virus due to the different heat and chemical conditions [19]. In addition, heat inactivation had a great impact on the amount of detectable RNA, which may cause false-negative results, especially in weakly positive cases [8].

Evidence has shown that a variety of commonly used disinfectants and laboratory inactivation procedures can reduce viral viability [17, 20]. This is especially important for healthcare settings including laboratories that require highly reliable inactivation methods to protect staff working with COVID-19 patients and samples [21].

The systematic study of the heat inactivation efficiency of rRT-PCR is still under investigation and validation remains to be studied and confirmed. Early detection has a significant impact on the prevention and control of SARS-CoV-2 samples. It is critical to identify individuals and facilitate the implementation of protective measures such as social distancing, quarantine, and isolation that help to mitigate the spread of the virus in the community.

Since many coronaviruses are heat sensitive, inactivation by heating samples to different temperatures prior to testing is considered a possible method to reduce the risk of transmission, but the effect of heating at different temperatures and for different time periods on the measurement of SARS-CoV-2 remains unclear. Thus, this study aims to evaluate the effect of heat inactivation at different temperatures and times and to determine the effect of chemical inactivation by bulk lysis on SARS-CoV-2 detection.

## Materials and Methods

### Study area and design

The laboratory-based experimental study design was conducted at EPHI National HIV Reference laboratory from August to November 2020 on the samples referred to the laboratory for COVID-19 testing. The National HIV reference laboratory is Ethiopia’s largest COVID-19 testing center, did perform COVID-19 testing since March 2019, and tested over 200,000 samples as of August 2020. The laboratory has a well-established quality system and is ISO 15189; 2012 accredited by Ethiopian National Accreditation Organization for HIV Viral load testing and early infant diagnosis. The study was conducted on all known SARS-CoV-2 samples with Known threshold cycle (CT) values.

### Sample size determination and sampling method

Eighty Nasopharyngeal/Oropharyngeal swab samples were selected and taken out from -80^0^C storage. The samples were stored with a disposable specimen collector containing 3-4 ml Viral Transport Media (VTM). These samples were collected between August 25/2020 to September 30/2020, from samples under investigation for SARS-CoV-2 and submitted to the EPHI, National HIV reference laboratory for diagnostic testing.

Positive samples with known Ct value were selected by simple random sampling technique, from 9,520 positive samples within one month, which is obtained from 28.4% prevalence of Covid-19 in Ethiopia as of August 20/2020. About 2380 samples were done in one week, then dividing this sample by 80 is 30, which was the interval number by which samples with known Ct values were selected. The first sample was selected by the lottery method.

### Specimen collection and testing

The nasopharyngeal/Oropharyngeal swab specimens were obtained according to CDC guidelines. All samples were registered with unique identification numbers (Barcode). All samples were tested with 2 controls (1 positive, 1 negative) when testing by reference method (Abbott Real-time SARS-COV-2 (EUA). Fig. 1 depicts the overall experimental procedure followed.

**Fig 1:**
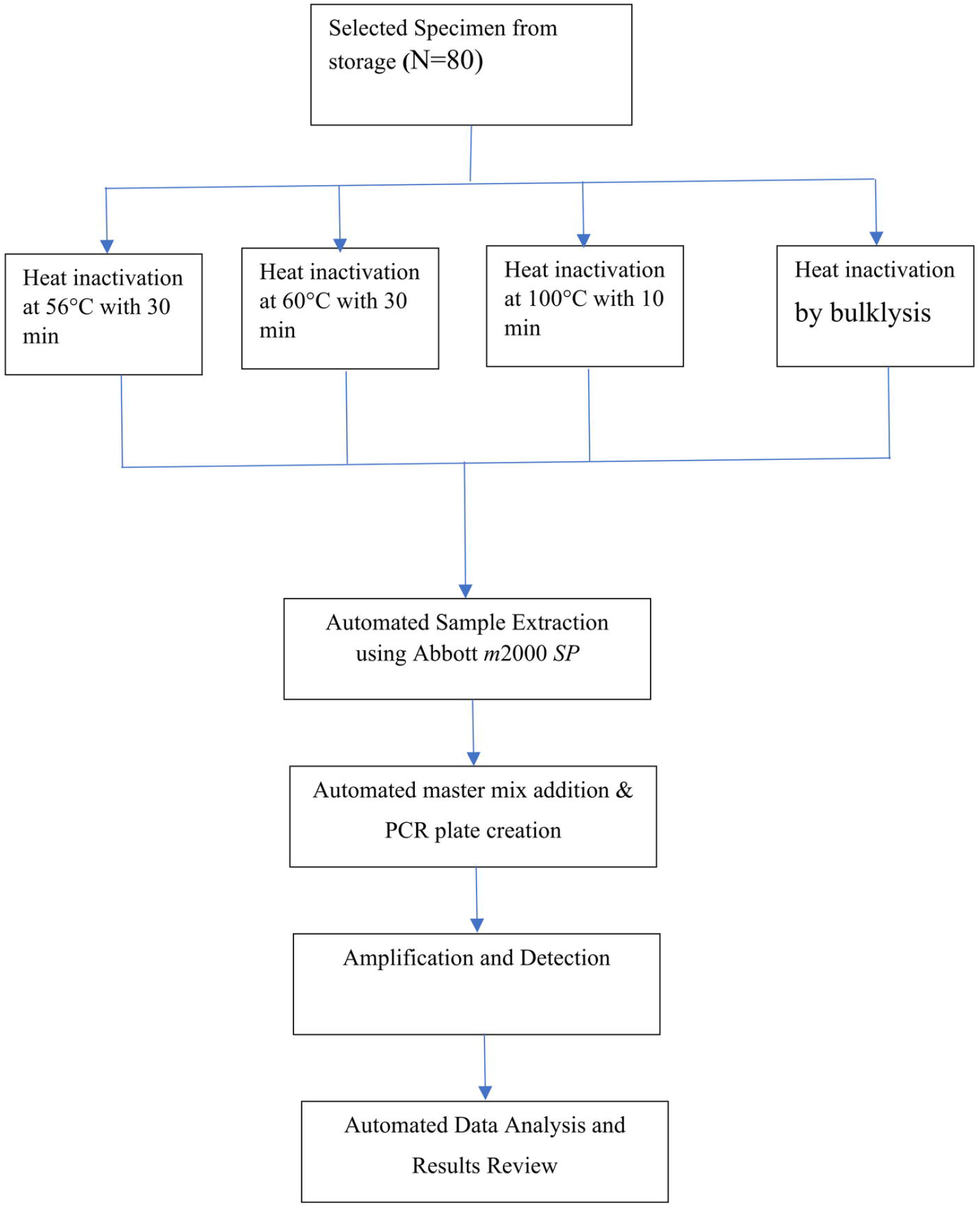
Process of specimen testing.

### Experiment

Tests were performed using the Abbott Real-time SARS-CoV-2 assay, a rRT-PCR test for the qualitative detection of SARS-CoV-2 in Nasopharyngeal and oropharyngeal swabs. Dual target assay for RNA-dependent RNA polymerase (RdRp) and N-genes detection of NAs from SARS-CoV-2 was analyzed. The assay also includes an internal control to indicate proper extraction, amplification, and detection. Results were reported as positive if the Ct value was <32, and defined as negative if the Ct value was 32 or more, based upon the signal threshold determined by the manufacturer [1].

The effects of heat treatment at different temperatures and durations and chemical inactivation on the SARS-CoV-2 rRT-PCR Ct-value were evaluated. The samples were inactivated at different temperatures in a water bath for different minutes (at 56°C for 30min, at 60°C for 30min, and at 100°C for 10min) and by chemical bulklysis. WHO recommends heat inactivation at 56°C for 30min and Abbott real-time RT-PCR is the golden method recommended for SARS-COV-2 detection and this temperature was the best for the viral inactivation [22-24]. After viral heat inactivation, NA extraction was done from 0.6 ml sample volume on the Abbott *m*2000*SP* instrument by using the Abbott *m*Sample Preparation System Deoxyribonucleic acid (DNA) according to the manufacturer’s recommendations [1]. On the other hand, Chemical inactivation was performed to see the effect of bulk lysis on the SARS-CoV-2 rRT-PCR. For chemical inactivation, bulk lysis was tested with the appropriate composition of sample and lysis proportion. As all samples were incubated with bulk lysis buffer and it was compared to the effect of heat inactivation methods. Each sample was incubated with the bulk lysis buffer at room temperature for 30 min; then, the sample was extracted by Abbott *m*2000*SP* instrument. After the extraction was completed, the samples of heat and chemical treated group was detected by Abbott m2000*rt* (12). The viral RNA was extracted from 500 μL of each sample, and the final elute was with 200 μL by using elution buffer [1, 22]. The amplification and detection of SARS-CoV-2 RNA was performed by Abbott *m*2000*rt* instrument targeted to dual-target assay for the RdRp and N genes [1]. The SARS-CoV-2 and internal control (IC) specific probes were each labeled with a different fluorophore (FAM™ (Carboxyfluorescein), ROX™, (Carboxy-X-rhodamine), and VIC® P (Proprietary dye) for target and IC detection, thus allowing for simultaneous detection of both amplified products. The test is a real-time RT-PCR test intended for the qualitative detection of NA from the SARS-CoV-2 in upper respiratory specimens [1].

### Data quality assurance

Samples were collected by well-trained professionals. All laboratory procedures were performed as per the documented standard operating procedures and according to specific manufacturing recommendations. Quality of each reagent like lot number, expiry date, storage conditions, physical leak-proof, no breakages, etc was checked before the actual laboratory analysis. Samples and reagents were stored at appropriate temperatures as indicated on the manufacturer inserts. Data was double-checked manually for completeness before data entry. We have used a respective recommended manufacturer protocol.

### Data Analysis

Data was analyzed and described by mean and standard deviation (mean±SD). Normality was assessed for the three temperature scenarios and bulk lysis, and they were violated normality. However, after we transformed by natural logarithm all of them were normally distributed. Repeated measurement analysis of variance was used to assess the mean difference between the three temperatures and bulk lysis on viral detection. To identify the place of significant difference post hoc analysis with modified Bonferroni correction was used.

## Results

### Reverse transcriptase real time polymerase chain reaction results of heat-inactivated samples at different temperatures and durations

The effect of heat treatment at different temperatures and durations on the SARS-CoV-2 rRT-PCR Ct-value was evaluated. All heat-inactivated samples at 56°C for 30min were tested positive. The Ct values of RdRp were 4.37–31.03 CN (cyclic number) at 56°C for 30min, 3.68– 30.64 CN at 60°C for 30min, and 3.37–28.74 CN at 100°C for 10min. The Ct values of the chemical bulklysis inactivated samples were from 3.62–27.74 CN except for those with weak positive samples as shown below in [Table 1] which were turned to negative results, as compared to the heat-inactivated samples at 56°C (WHO recommended Standard temperature for heat inactivation of SARS-COV-2, the cut-off Ct value for Abbott m2000*rt* machine is 32). Heat inactivation methods resulted in the reduction of positive SARS-CoV-2 samples to undetectable levels, especially in weak positive samples. About 6.2% (5/80) of samples were changed to negative results in heat inactivation at 60°C and 8.7% (7/80) of samples were changed to negative in heat inactivation at 100°C. The heat inactivation at 100°C for 10 mins for those with high Ct value (low viral load concentration (weak positive samples)) was changed to a negative. Comparison of heat inactivation of weakly positive samples at 56°C with heat inactivation at 60°C, 100°C, and chemical bulklysis were summarized in percentage as shown below. [Table 1]

**Table 1.** Cycle threshold values for RdRp measured from swab samples following heat treatments at different temperatures and chemical bulklysis EPHI, Ethiopia.

| S. N | Ct value results |  |  |  |
| --- | --- | --- | --- | --- |
|  | T1 | T2 | T3 | Bulklysis |
| 1 | 30.11 CN | Negative | Negative | Negative |
| 2 | 31.03 CN | Negative | Negative | Negative |
| 3 | 28.57 CN | Negative | Negative | Negative |
| 4 | 27.13 CN | 28.37 CN | Negative | 27.74 CN |
| 5 | 26.22 CN | 30.64 CN | Negative | 26.62 CN |
| 6 | 29.65 CN | Negative | Negative | 28.89 CN |
| 7 | 30.54 CN | Negative | Negative | 28.99 CN |
| % |  | 6.2% | 8.7% | 3.7% |

T1=Temperature at 56°C, T2=Temperature at 60°C, T3=Temperature at 100°C, CN=Cyclic number, Ct=Cyclic threshold

The Ct values of heat-inactivated samples (at 60°C, 100°C) and bulk lysis were significantly different from the temperature at 56°C (as compared to WHO heat inactivation standard temperature at 56°C) (*p*=0.01, *p*=0.001) and *p*=0.02 respectively. The place of significant difference was identified by using post hoc analysis with modified Bonferroni correction. [Table 2]

**Table 2.**
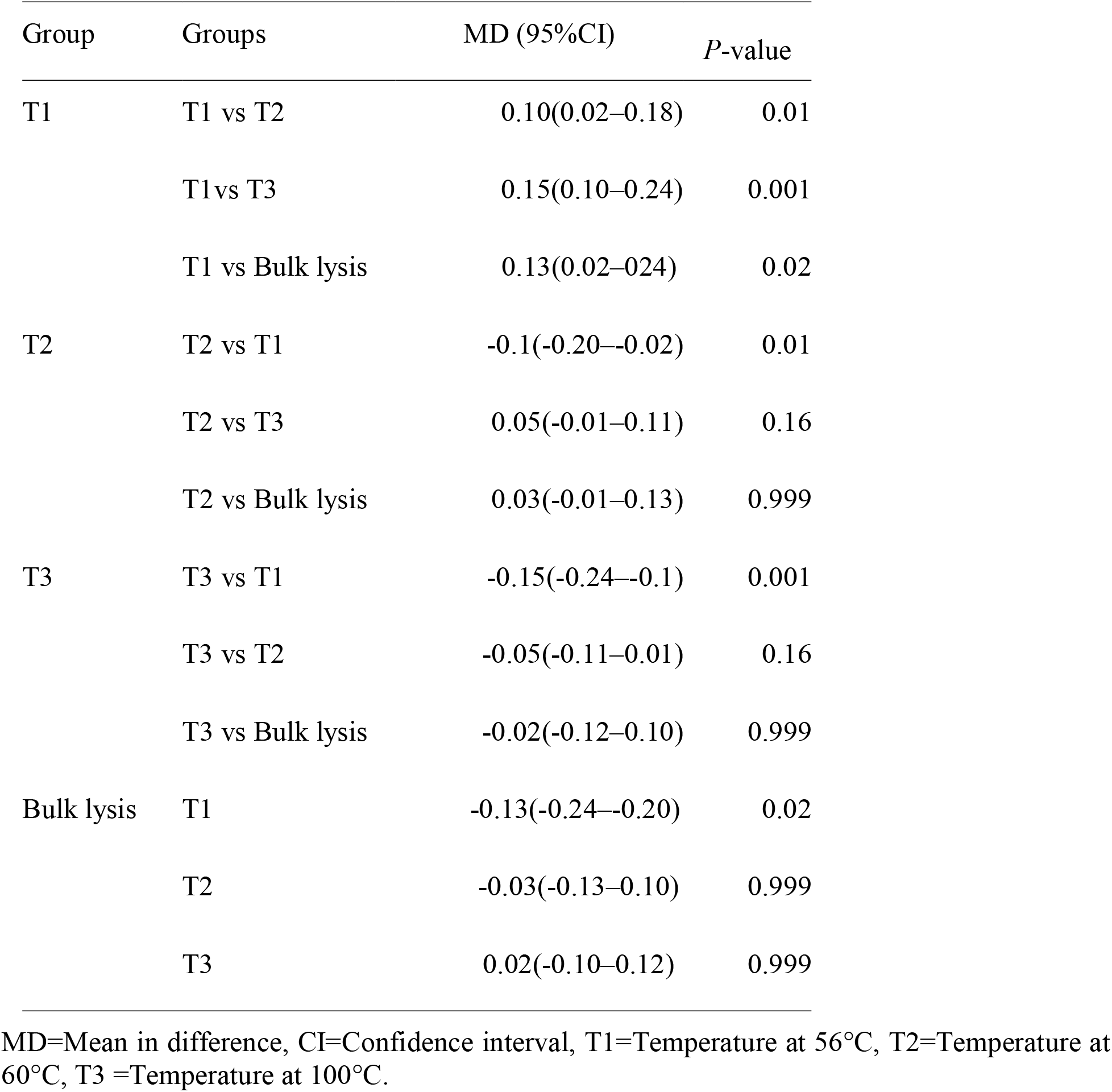
Mean difference of group with their confidence interval and *p*-value following heat inactivation at different temperatures and durations EPHI, Ethiopia.

### Heat inactivation by chemical bulk lysis

All samples were incubated with chemical bulk lysis buffer, (as it was compared with AVL buffer, AVL serves as the standard for the comparison of the diverse chemical inactivation methods). The decline in the viral RNA quantity was observed in some of the samples treated with chemical bulklysis, especially for those with high Ct values stored at room temperature for 30 mins. It was shown that 3.7% (3/80) of analyzed samples were turned to negative (from weak positive samples). There were significant differences between the Ct values of bulklysis and heat inactivation at 56°C for the RdRp genes (Repeated measure ANOVA; *P* = 0.02). Following the comparison of bulk lysis to all other forms of inactivation used in this study, only heat-inactivated at 56°C was significantly different from bulklysis inactivation for the RdRp genes.

The mean average Ct values of the heat-treated samples at different temperatures and chemical bulklysis for different durations (at 56°C for 30 min, 60°C for 30 min, 100°C for 10 min, and bulklysis) as shown below by bar (Fig 2).

**Fig 2:**
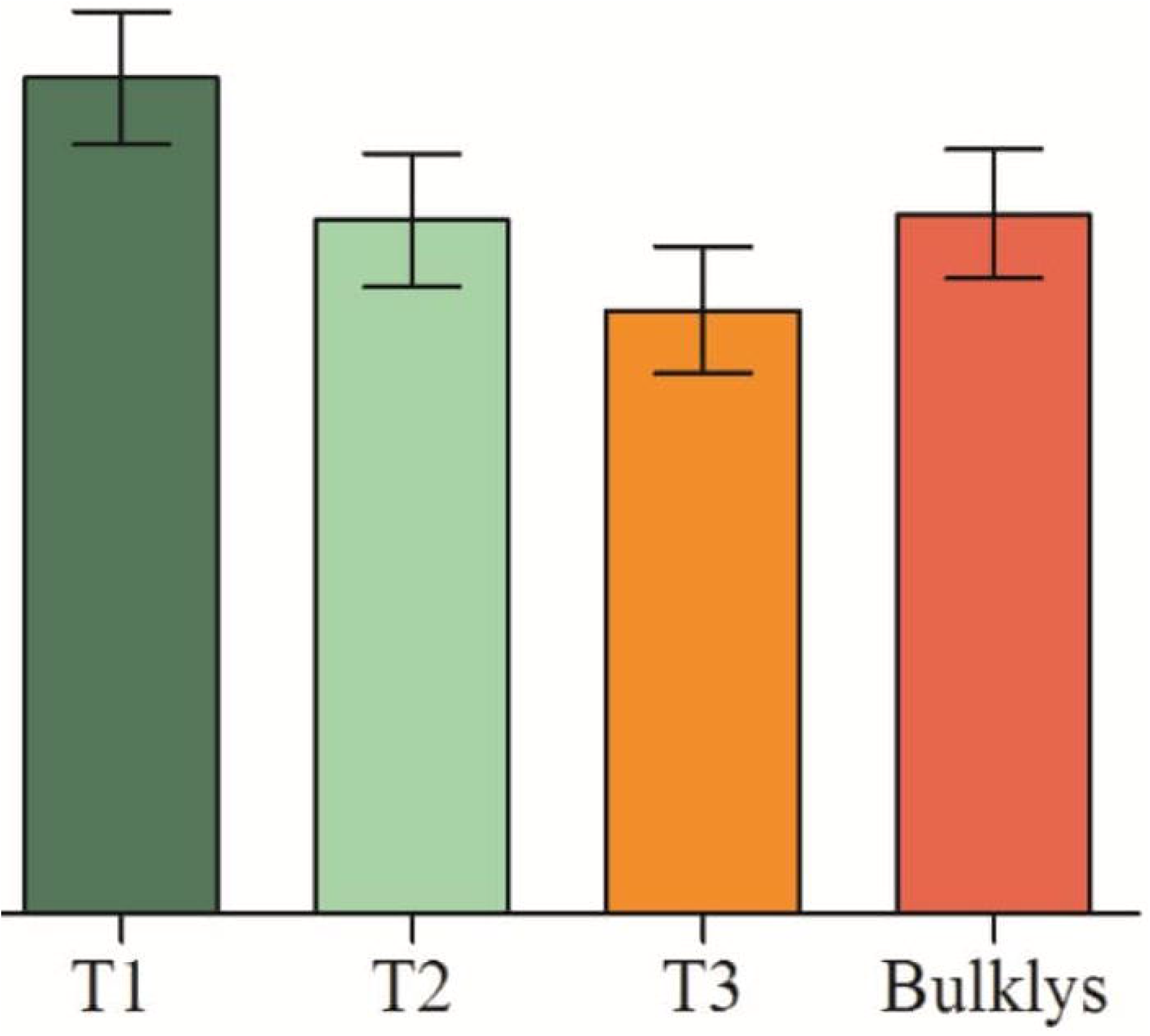
Mean cyclic threshold comparison between the three temperatures and bulklysis. (The standard errors bar shows mean ±SD): T1-Temperature1= at 56°C; T2-temperature 2= at 60°C and T3-temperature 3= at 100°C; SD-standard deviation).

## Discussion

SARS-CoV-2 has been emerged in late 2019 in China and now is on the list of Public Health Emergency of International Concern (PHEIC). Respiratory specimens have been used to diagnose SARS-CoV-2 infection by Abbott rRT-PCR, and are regarded as the main detection method and the gold standard. Following the rapid global spread of SARS-CoV-2 and the need for universal testing, more and more individuals are exposed to non-inactivated virus samples, thereby increasing the chances of occupational infection. The WHO and United States CDC have released laboratory guidelines to mitigate the risk of exposure during diagnostic and research procedures [25, 26, 27]. Despite recommendations for handling within contained biosafety cabinets, individuals working with these samples are still required to handle potentially infectious viruses at the initial steps of acquiring and preparing samples before testing, thereby increasing the risk of exposure. Therefore, the continued need for COVID-19 testing worldwide requires the utilization of simple and effective inactivation techniques.

It has been shown that in the SARS-CoV-2 swab sample, the amount of SARS-CoV-2 could be reduced at 60°C for 30min and bulklysis, but still infectious. Only heating at a temperature of 100°C for 10min was able to inactivate it totally, high temperature with high duration [12]. However, we have been trying to demonstrate that it is possible to ensure the test integrity by applying heat inactivation at several conditions. RT-PCR Ct values are defined as the number of cycles of amplification required for the accumulated fluorescence (produced by target gene amplification) to reach a threshold above the background. Ct values are therefore inversely related to viral load; low Ct values indicate high viral loads and high Ct values indicate low virus NA concentration in the sample. With regards to SARS-CoV-2, low Ct values from SARS-CoV-2 samples have been reported to correlate with an increased probability of developing severe disease and mortality [28]. In this study, the Ct value was significantly affected by heating at 60 °C and 100 °C for those with weak positive samples. This is in agreement with the studies of Pastorino, (2020) [12]. Therefore, an increment in Ct value may lead to a reduction in RT-PCR sensitivity that could have an impact on clinical diagnosis. The less notable increase in Ct value observed when the virus was heated to 100 °C, could be attributed to the shorter heating time, this is in line with the study by Zou *et al*., 2020 [25]. Lower temperature heat treatment combined with chemical inactivation, short-duration high-temperature heat-treatments, or chemical inactivation alone may be more appropriate to preserve RNA integrity and maximize PCR optimization for detection of SARS-COV-2 RNA from low-concentration SARS-COV-2 samples. Our results show significant variation in the effect of heat-treatment inactivation on the SARS-CoV-2 detection. This emphasizes the importance of local validation of inactivation methods and the need for consistency in inactivation protocols to ensure sufficient reduction in virus concentration for the processing of clinical samples.

Weak positive samples may become false negatives in SARS-CoV-2 RT-PCR detection. Our study also has shown that the heat-inactivated samples at 56°C were consistent with those in heat-inactivated ones at 60°C, 100°C, and chemical bulk lysis for low CT value results, which agrees with the study done by Pastorino, B., 2020 and Pan, Y., 2020 [12, 29]. Heating at 100□ for 10min would result in a false negative, which is consistent with that of heating at 92□ for 15min, the SARS-CoV-2 RNA copies in a sample were dropped significantly as the study done by Zou *et al*., 2020 and Burton, J., 2021[25, 30]. In fact, studies have suggested that the sample cells shall die and rupture in the event of relatively high temperature and long durations, leading to the release of a large number of cell nucleases, and then a large amount of RNA degradation, which may contribute to false negatives in NA detection. We hypothesize that heating at 100□ for 30 min or 60 min (high temperature for a long time) lyse a large number of cells, and expose RNA to RNases present in the samples, and the duration is also a key factor.

Although our study showed that heating at 100□ for 10min was consistent with heated one’s at 56°C, especially for those with strong positive samples, except for those samples with weak positive, strong positive samples were showed a tendency to decrease in Ct values to some extent, this is in agreement with the study done by Wang, T., 2020 [31]. The RNA preservation may be due to the preservation chemical, which contains guanidine isothiocyanate. This suggested that the presence of the preservation chemical can effectively protect the integrity of the viral nucleic acid, thereby increasing the proportion of detectable nucleic acids. It may also be that we used the original samples rather than the diluted samples. However, many factors can influence the effect of a lysis buffer, including the virus concentration, nature of sample matrix, contact time and reaction temperature, virus structure, and concentration/composition of the lysis buffer (denaturing agent and/or detergent, pH) used.

### Ethical Clearance

Ethical clearance was obtained from EPHI Scientific ethical review committee. Approval and official permission was obtained from the EPHI research committee to use the leftover samples in the laboratory (EPHI-IRB-296-2020). The confidentiality of the data collected was kept to a maximum and each patient identity was coded.

### Limitation of the study

Performance has only been established with the specimen types listed in the Intended Use. Other specimen types have not been evaluated and should not be used with this assay.

The small sample size can affect the conclusion.

Since our study conducted analysis from stored positive samples, weak positive samples may be affected by repeated thaw and refreezing.

## Conclusion and Recommendations

We found that the effect of heat-inactivation varies greatly depending on temperature and duration. The impact of chosen inactivation method on the sensitivity of subsequent SARS-CoV-2 detection should be assessed locally. The effect of heat on Ct value should be considered when interpreting diagnostic PCR results from clinical samples which could have an initial low virus concentration. Since high temperature and chemical bulklysis have the tendency to release a large number of RNA and used it for sequencing purposes (preserve and optimize). Our findings need to be further verified by other large-scale research and virus infectivity experiments

## Data Availability

All data produced in the present work are contained in the manuscript

## List of Abbreviations

CDC: Center Disease Control and Prevention
CT: Cyclic Threshold
CN: Cyclic Number
DNA: Deoxyribose nucleic acid
EPHI: Ethiopian Public Health Institute
EUA: Emergency Use Authorization
IC: Internal control
ISO: International Standard for Organization
SARS-COV-2: Severe Acute Respiratory Syndrome Coronavirus 2
RNA: Ribonucleic acid
WHO: World Health Organization
RT-PCR: Real-time Polymerase Chain Reaction
RdRp: RNA dependent RNA polymerase
VTM: Viral Transport Media

## Facilities available for the study

The study was conducted in the Ethiopian public health institute. We were also, using suitable data storage devices and a recent version of statistical packages that are available at EPHI for data storage and analysis.

## Authors’ contributions

DL has participated in the design, coordination of the study, and data collection, analysis, and interpretation. GG has contributed in, drafting, revising, or critically reviewing the article and revision and amendment of the draft manuscript. GGH has contributed in wright up, analysis and review, RD has contributed in wright up, analysis, and review, GB has contributed in data collection and analysis, TS has contributed in data collection and analysis, DA has contributed in data collection and analysis, DC has contributed in data collection, analysis, and manuscript preparation, BL has contributed in wright up, analysis and review, JB has contributed to conceptualization and methodology, SA has contributed to conceptualization and writeup, HHT has contributed to the design, including final approval of the version to be published. All authors read and approved the final manuscript.

## Acknowledgments

We are grateful in extending our thanks to Ethiopian Public Health Institute for allowing as the setup, ethically approve our research and allow as to use the leftover samples. We also extend our appreciation to Mr. Atsbeha G/eigziebxier for his unreserved contribution.

## Supporting Information

S1 Fig 1: Process of specimen testing

S2 Fig2: Mean cyclic threshold comparison between the three temperatures and bulklysis

S1 Table1: Cycle threshold values for RdRp measured from swab samples following heat treatments at different temperatures and chemical bulklysis.

S2 Table2: Mean difference of group with their confidence interval and p-value following heat inactivation at different temperatures and durations.

